# Timing, safety, and efficacy of initiating anticoagulation in intracranial surgery: a systematic review and meta-analysis

**DOI:** 10.1101/2025.11.07.25339766

**Authors:** Farzan Fahim, Negin Safari Dehnavi, Farbod Tabasi Kakhki, Parniya Amini, Reza Saeedinia, Mohammad Maroufi, Ali Saravani, Mana Majlesi, Ahmad Fathinejad, Alireza Zali, Saeed Oraee Yazdani

## Abstract

**Background:** Venous thromboembolism (VTE) remains a major cause of early postoperative morbidity and mortality after intracranial tumor surgery, yet consensus on the optimal timing of pharmacologic prophylaxis is lacking.

**Methods:** We conducted a PRISMA-guided systematic review and meta-analysis of randomized and cohort studies evaluating postoperative anticoagulant prophylaxis after brain tumor craniotomy. PubMed, Embase, Web of Science, Scopus, and Cochrane were searched through September 9, 2025. Two reviewers independently screened studies and extracted data; risk of bias was evaluated using JBI tools. Primary and secondary outcomes were VTE, intracranial hemorrhage (ICH), and mortality. Odds ratios (ORs) were pooled using random-effects (REML) models with Hartung–Knapp adjustment. Prespecified sensitivity analyses; leave-one-out, LMWH-only, postoperative-only, UFH vs LMWH subgrouping, and common- vs random-effects models, assessed robustness.

**Results:** Six studies (seven comparisons) met inclusion criteria. Pharmacologic prophylaxis demonstrated a directionally favorable but non-significant reduction in VTE (OR 0.71, 95% CI 0.35–1.45; I^2^=28.9%), with no increase in ICH (OR 0.96, 95% CI 0.46–2.00; I^2^=0%) or mortality (OR 0.74, 95% CI 0.33–1.65; I^2^=0%). Sensitivity analyses showed strong internal consistency: excluding a mechanistically atypical UFH-based hemodilution study (Moussa 2016) removed all heterogeneity (I^2^=0%) and yielded pooled effects nearly identical to LMWH-only and postoperative-only models (ORs 0.89–0.92). Apparent funnel-plot asymmetry was resolved upon exclusion of this structural outlier, indicating no meaningful publication bias.

**Conclusions:** Across available evidence, initiating LMWH prophylaxis 12–72 hours after tumor craniotomy appears to optimize the balance between thrombosis prevention and hemorrhagic safety. Preoperative initiation provides no clear benefit and may increase bleeding risk. Larger, standardized trials are needed to refine timing, dosing, and patient selection.

## Introduction

Venous thromboembolism (VTE), encompassing deep vein thrombosis (DVT) and pulmonary embolism (PE), is among the most consequential complications after neurosurgical procedures (1–3). Its clinical significance derives from substantial morbidity and the disproportionate risk of early death; PE alone accounts for up to 20–30% of postoperative mortality in this setting (4–6). Patients with intracranial tumors face a particularly elevated risk because of tumor-driven hypercoagulability, endothelial injury from surgery, and postoperative immobility (7–9). Within this group, meningioma surgery carries a distinctly high burden. In a retrospective cohort study, VTE occurred in 15.4% of patients managed with mechanical prophylaxis alone, whereas it was absent in those receiving a combination of low-molecular-weight heparin (LMWH) and hemodilution (10). A population-based study conducted in Scandinavia indicated VTE rates of 3–4% following meningioma resection, with mortality related to PE reaching 23% (11). All of these observations show that patients who have meningioma surgery are at a higher risk of thromboembolism than other patients.

Multiple strategies have been evaluated to mitigate this risk. Mechanical measures such as elastic stockings and intermittent pneumatic compression (IPC) are widely adopted, yet they are often insufficient for high-risk patients when used in isolation (12–14). Pharmacologic prophylaxis with unfractionated heparin (UFH) or LMWH has therefore been explored. A prospective randomized trial demonstrated that perioperative minidose heparin did not significantly increase bleeding compared with placebo (15). By contrast, initiating LMWH preoperatively was associated with a 10.9% rate of clinically significant postoperative intracranial hemorrhage, prompting early termination (16). Later studies have shown that starting anticoagulation after surgery, typically within 24 to 72 hours, reduces the risk of thromboembolic events without increasing the risk of clinically significant hematomas (17, 18).

Despite these advances, the optimal timing and method of prophylaxis remain unclear. Established risk factors, such as advanced age, increased tumor size, skull-base location, and delayed mobilization, further complicate individualized decision-making (10). Multimodal strategies integrating mechanical prophylaxis, timely anticoagulation, and hydration seem to be the most effective; however, a universally accepted standard has yet to be established (11, 19, 20). This systematic review and meta-analysis seek to delineate the incidence of VTE following meningioma resection, evaluate the efficacy and safety of mechanical, pharmacologic, and combined prophylaxis strategies, and elucidate the impact of early versus delayed anticoagulation on thromboembolic protection and hemorrhagic risk.

## Methods

### Registration, protocol, and reporting

This systematic review followed the PRISMA 2020 guidelines (21, 22) The protocol was registered in PROSPERO prior to screening (CRD420251157525). The PRISMA checklist, the full protocol, the exact database search strings, and the data-extraction template are provided in the Supplementary Materials (Appendix 1-4).

### Information sources and search strategy

We searched PubMed, Embase, Web of Science Core Collection, Scopus, and the Cochrane Library from database inception through 9 September 2025. No date or language restrictions were applied. Searches combined controlled vocabulary and free-text terms for intracranial tumor surgery, thromboembolic or hemorrhagic outcomes, pharmacologic prophylaxis, and timing. The exact strategies implemented in each database are reproduced verbatim in the Supplementary Materials.

This is an example of pubmed search strategy:

(craniotom* OR craniectom* OR “skull base surg*“[tiab] OR “transsphenoidal surg*“[tiab] OR “awake craniotomy“[tiab] OR “burr hole*“[tiab] OR “microvascular decompression“[tiab] OR hemispherect*[tiab]) AND (“Brain Neoplasm“[ tiab] OR “Pituitary Neoplasm“[tiab] OR “Skull Base Neoplasm“[tiab] OR “brain tumor“[tiab] OR “brain tumour“[tiab] OR “intracranial tumor*“[tiab] OR “intracranial tumour*“[tiab] OR “glioma“[tiab] OR “glioblastoma“[tiab] OR “meningioma“[tiab] OR “astrocytoma“[tiab] OR “pituitary adenoma“[tiab]) AND (“Hemorrhage“[tiab] OR bleeding[tiab] OR thrombo-embolism[tiab] OR Thromboembolism [tiab] OR “coagulation“[tiab] OR Complication[tiab] OR “Venous Thrombosis“[MESH] OR “Pulmonary Embolism” [tiab] OR thromboprophylaxis[tiab] OR chemoprophylaxis[tiab] OR “VTE prophylaxis“[tiab] OR “DVT prophylaxis“[tiab] OR anticoagula*[tiab] OR heparin[tiab] OR “low molecular weight heparin“[tiab] OR enoxaparin[tiab] OR dalteparin[tiab] OR tinzaparin[tiab] OR fondaparinux[tiab]) AND (timing[tiab] OR time[tiab] OR early[tiab] OR delayed[tiab] OR delay[tiab] OR hour*[tiab] OR hr[tiab] OR hrs[tiab] OR “24 h“[tiab] OR “24h“[tiab] OR “within 24“[tiab] OR “first 24“[tiab] OR “after 24“[tiab] OR “postoperative day 1“[tiab] OR “day 1“[tiab])

The complete database search strategies are provided in Appendix2.

### Eligibility criteria

We included studies enrolling patients who underwent intracranial tumor surgery of any type, in which prophylactic anticoagulant therapy was administered after surgery. Eligible interventions comprised antithrombotic prophylaxis with anticoagulant or antiplatelet agents intended to prevent DVT or VTE, with comparators reflecting different initiation windows, such as within 24 hours versus 24–48 hours postoperatively. Where applicable, studies evaluating mechanical prophylaxis at varying postoperative time points were also considered.

The primary outcome was postoperative intracranial hemorrhage at the resection site, confirmed by imaging. Secondary outcomes were the incidence of DVT and PE as measures of prophylactic efficacy, and rebleeding as the principal secondary safety outcome.

Eligible designs were randomized trials and prospective or retrospective cohort studies. Case series with two or more patients were considered for qualitative description and were included in quantitative synthesis only when variance data were available or could be derived. We excluded in vitro or animal studies, studies unrelated to tumor surgery, studies without postoperative prophylactic drug exposure, studies lacking a comparison group, abstracts-only records, reviews, editorials, and duplicate reports.

### Study selection

Two reviewers screened titles and abstracts in duplicate, followed by an independent full-text review. Specifically, AS and MM completed title and abstract screening; AS and PA performed full-text assessments; NS resolved disagreements. The PRISMA flow diagram summarizes screening and inclusion: 6,232 records identified; 3,775 removed before screening; 2,457 screened at the title and abstract level; and 2,354 exclusions for reasons including not related, case report, review, duplicate, case series, or animal study. All potentially eligible full texts were retrieved; six studies met the inclusion criteria and were included in the review and meta-analysis.

### Risk of bias assessment

Methodological quality was appraised using the Joanna Briggs Institute Critical Appraisal Checklists tailored to study design. AF and MM conducted independent assessments with adjudication by NS. For each study, we calculated the JBI percentage score and assigned an overall risk category: low risk (80 to 100 percent), moderate risk (50 to 79 percent), or high risk (20 to 49 percent). Domains included clarity of inclusion criteria, appropriateness of sampling and recruitment, baseline similarity of groups when applicable, validity and reliability of exposure and outcome measurement, identification and control of confounders, completeness of follow-up, and appropriateness of statistical analysis. Item-level results are provided in the Supplementary Materials.

### Data extraction

Using a piloted standardized form, PA and RS independently extracted study identifiers and metadata, funding source, country or region, design, sample size, whether analyses were adjusted or unadjusted, center type, period of data collection, inclusion and exclusion criteria, follow-up duration, ethics approval, conflicts of interest, and the main findings. Discrepancies were resolved by consensus.

### Statistical analysis

All analyses were carried out in R version 4.5.1 using the updated meta package. Odds ratios and 95 percent confidence intervals were pooled with a random-effects model using restricted maximum likelihood for the between-study variance. Confidence limits for random-effects estimates were computed with the Hartung–Knapp adjustment. Heterogeneity was assessed with the I^2^ statistic, the between-study variance, and Cochran’s Q test. Subgroup analyses were planned a priori to evaluate the timing of prophylaxis initiation, comparing preoperative administration with postoperative initiation between 12 and 72 hours.

Potential small-study effects and publication bias were visually evaluated using funnel plots generated separately for each endpoint. For venous thromboembolism, five studies were distributed around the central line, with approximately log odds ratios of 0.7, a slight predominance on the right, and overall symmetry. For DVT, six studies appeared largely symmetric, with two smaller studies along the left boundary at log odds ratios of approximately 0.02 to 0.1 and standard errors near 1.5, a pattern consistent with minimal asymmetry. For intracranial hemorrhage, the plot was nearly symmetric around a log odds ratio of 1.0, except for two minor, imprecise high-odds-ratio points near a standard error of 1.5, reflecting rare hemorrhage events. For mortality, four studies were evenly distributed within the funnel centered near a log odds ratio of 0.7 without visual evidence of asymmetry. Across outcomes, there was no convincing graphical evidence of publication bias.

### Sensitivity and Robustness Analyses

To evaluate the robustness of the pooled estimates, we performed a predefined series of sensitivity analyses in accordance with PRISMA 2020 recommendations. These included:

1. a leave-one-out influence analysis in which each study was iteratively removed to identify excessively influential estimates;
2. a reanalysis excluding the Moussa et al. study, which used unfractionated heparin and combined intraoperative hemodilution, representing a mechanistically distinct and clinically atypical regimen compared with the LMWH-based protocols used in all other trials;
3. a postoperative-only sensitivity model excluding studies initiating prophylaxis preoperatively;
4. a subgroup analysis stratified by pharmacological class (LMWH vs UFH); and
5. replication of pooled effects under both common-effect and random-effects (REML, Hartung–Knapp adjustment) models.

All sensitivity models used the same effect size metric (Odds Ratio) and identical computational framework as the main analysis. Funnel plots were visually inspected in both the full dataset and after exclusion of the Moussa et al. study to evaluate small-study asymmetry while avoiding misinterpretation in the presence of an outlier.

## Results

### Study selection

The search yielded 6,232 records (Scopus: 2,973; Embase: 1,375; Web of Science: 1,219; PubMed: 589; Cochrane: 76). After removing 3,775 duplicates, 2,457 titles and abstracts were screened, and 2,354 were excluded as not relevant to the topic, case reports, reviews, duplicates, case series, or animal studies. We retrieved 103 full-text reports; 97 were excluded because of drug incompatibility, incomplete articles, non-tumor surgery, outcomes not related to VTE, absence of a comparison group, or no intervention. Six unique studies met the eligibility criteria and were included in the quantitative synthesis. One study reported DVT and PE separately, yielding seven total comparisons. The study selection process is summarized in Figure 1 (PRISMA flow diagram).

**Figure 1.**
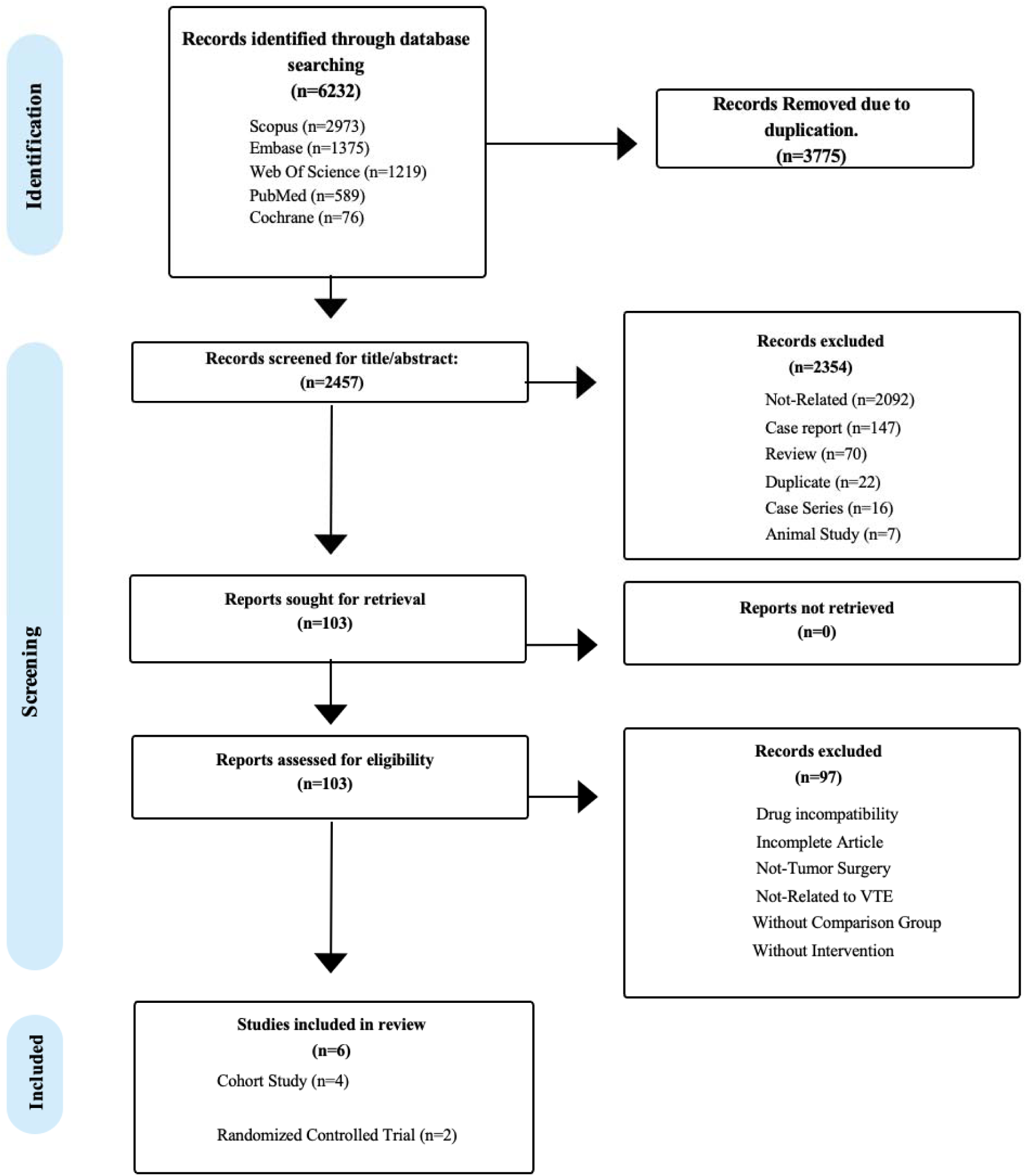
PRISMA 2020 flow diagram of study selection.

### Study characteristics

The six studies evaluated pharmacologic thromboprophylaxis after intracranial tumor surgery compared with no prophylaxis. Study designs included four cohort studies and two randomized controlled trials. Timing of anticoagulation initiation varied across studies, with preoperative regimens and postoperative starts ranging from 12 to 72 hours. Types of heparin used included LMWH and UFH. (Table 1)

**Table 1.** Overview of included studies.

| Authors-Year | Country-Region | Study design | Data Collection Period | Center-Type | Founding source |
| --- | --- | --- | --- | --- | --- |
| Wael Mohamed Mohamed Moussa, Mohamed Abbas, Aly Mohamed, 2016 | Egypt | retrospective cohort | 1st of February 2010 - 31st of January 2015 | single-center study (Department of Neurosurgery, Faculty of Medicine, Alexandria University, Alexandria, Egypt) | no external funding source |
| Lawrence D. Dickinson, M.D., Lisa D. Miller, R.N., Chirag P. Patel, B.S., Sanjay K. Gupta, M.D.-1998 | USA | Prospective Randomized Unblinded Clinical Trial | a period of 20 months | single-center study, conducted exclusively at the University of Michigan Hospital, Department of Neurosurgery, Ann Arbor, Michigan. | no external funding source |
| SHLOMI CONSTANTINI, M.D., M.SC., ANDY KANNER, M.D., ADI FRIEDMAN, M.D., YGAL SHOSHAN, M.D., ZVI ISRAEL, M.D., ELI ASHKENAZI, M.D., MOSHE GERTEL, M.D., AMIRAN EVEN, PH.D., YOSHI SHEVACH, M.D., MORDECHAY SHALIT, M.D., FELIX UMANSKY, M.D., AND ZVI H. RAPPAPORT, M.D.-2001 | Israel | RCT (prospective, double-blind) | a period of 14 months | multicenter, conducted in academic, university-affiliated medical centers in Israel | no external funding source |
| Tene A. Cage Æ Kathleen R. Lamborn Æ Marcus L. Ware Æ Anna Frankfurt Æ Lenna Chakalian Æ Mitchell S. Berger Æ Michael W. McDermott-2009 | USA | retrospective cohort | 2000-2005 | single-center study - University of California, San Francisco (UCSF) Medical Center. | no external funding source |
| Kristin Sjaavik, Jiri Bartek J, Ole Solheim, Tor Ingebrigtsen, Sasha Gulati, Lisa Millgard Sagberg, Petter Fjorander Asgeir Store Jakola-2016 | Scandinavia | retrospective, cohort | January 1, 2007 - June 30, 2013 | multicenter, Department of Neurosurgery, University Hospital of North Norway, Tromsø, Norway; Department of Neurosurgery, Karolinska University Hospital, Stockholm, Sweden; Department of Neurosurgery, St. Olavs University Hospital, Trondheim, Norway | no external funding source |
| Robert G. Briggs, Yueh-Hsin Linb, Nicholas B. Dadarioe, Isabella M. Youngc, Andrew K. Connerd, Wenjai Xud, Onur Tanglayb, Sihyong J. | Australia | retrospective cohort | 2012-2017 | single center - Prince of Wales Private Hospital, Randwick, New South Wales, Australia. | no external funding source |
| Kimb, R. Dineth Fonsekab<br>Phillip A. Bonneyd, Arpan<br>R. Chakrabortyd Cameron<br>E. Nixd, Lyke R.<br>Flecherd,Jacky T. Yeungb,<br>Charles Teob, Michael E.<br>Sughrueb-2021 |  |  |  |  |  |

A comprehensive summary of the extracted data from the included studies is provided in Table 2.

**Table 2.** Summary of extracted data of included studies.

| study | Intervention Drug | Dose | Dose interval | Timing_of_Initiation hours | Duration_of Prophylaxis | Mechanical Prophylaxis | Follow-up Duration |
| --- | --- | --- | --- | --- | --- | --- | --- |
| Moussa, 2016 | Enoxaparin | 40mg subcutaneously | every 24 h | 12hours after surgery | Until full ambulation | Elastic stockings (both groups),No pneumatic compression , Early mobilization (both groups) | Mean = 150 days<br>Range = 30 – 390 days |
| Dickinson, 1998 | Enoxaparin | 30 mg subcutaneously | every 12 h | Pre-anesthesia induction | until discharge | Thigh-high SCDs (Kendall) | 1 month postoperatively |
| Constantini, 2001 | heparin | subcutaneously | every 12 h | 2 hours before surgery | Until full ambulation | Not Reported | 72 hours postoperatively |
| Cage, 2009 | Enoxaparin | 40mg subcutaneously | every 24 h | within 24–48 hours after surgery | ranged from one to seven days | TED hose stockings and sequential compression devices,all patients) | 30 days postoperatively |
| Sjävik, 2016 | Enoxaparin | 40mg subcutaneously | every 24 h | 12 hours before surgery | Until sufficiently mobilized | compression stockings or SCD,all patients | 30 days postoperatively |
| Briggs, 2021 | Enoxaparin | 40mg subcutaneously | every 24 h | within 24– 72 hours after surgery | minimum of 30 days, unless hemorrhagic complications occurred | (SCDs) during hospital stay | reviewed through the last available follow-up |

### Risk of bias

Methodological quality was assessed using the Joanna Briggs Institute (JBI) Critical Appraisal Checklists, applied according to the study design and rated independently by two reviewers, with adjudication by a third. Five studies were judged to have a medium (moderate) risk of bias, and one study was rated low risk; none were high risk. JBI percentage scores ranged from 53.8% to 81.8%, with a median of approximately 68%. The principal concerns relate to selection and recruitment procedures and to the handling of confounding and co-interventions in observational designs, with occasional incomplete follow-up reporting. Outcome measurement was generally sound. The domain-level matrix and study-level summaries are shown in Figure 2 and Table 3.

**Figure 2.**
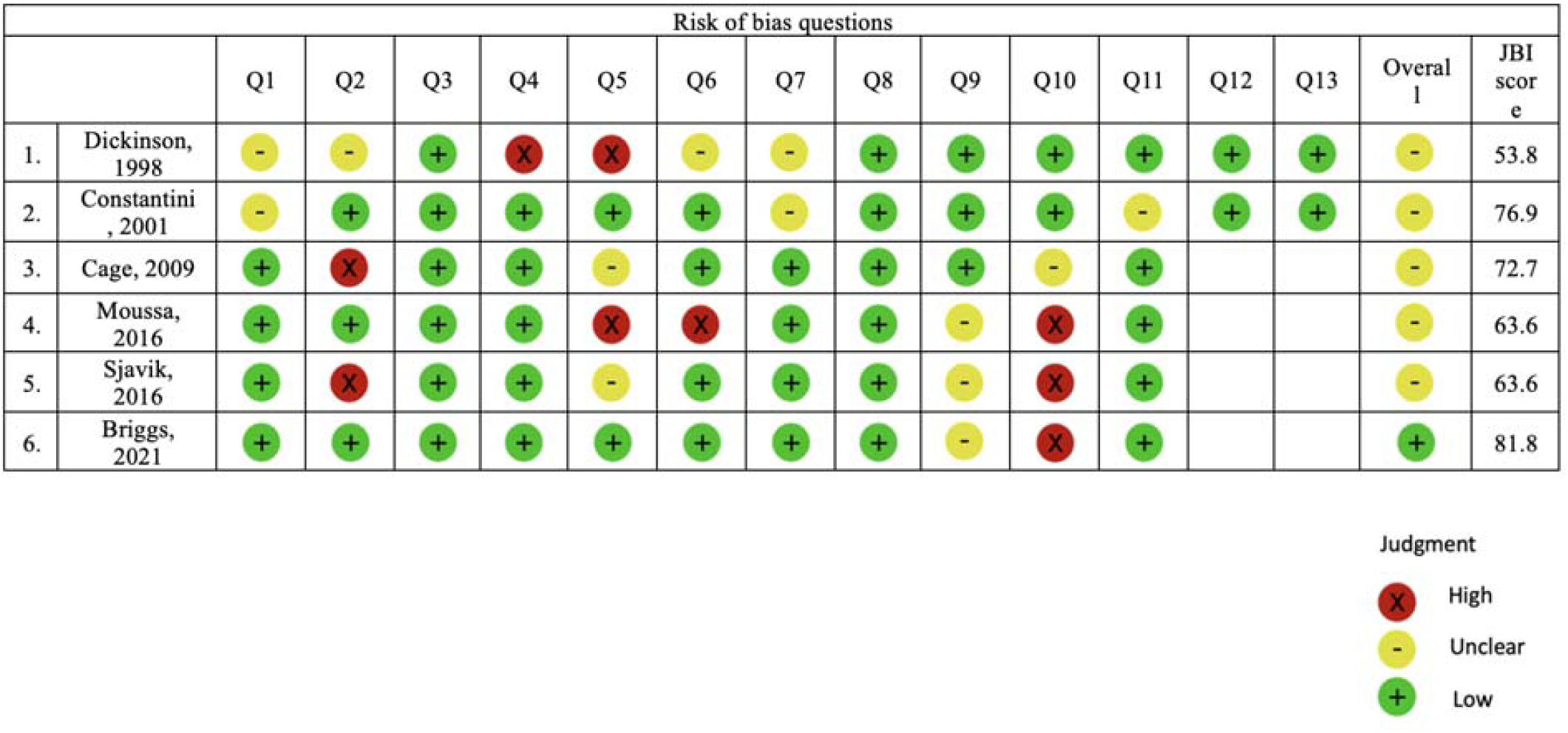
Risk of bias assessment using the Joanna Briggs Institute (JBI) critical appraisal tools.

**Table 3.** Risk of Bias Assessment of Included Studies.

| Study | Study Design | Random Sequence Generation | Allocation Concealment | Blinding of Participants / Assessors | Incomplete Outcome Data | Selective Reporting | Overall Risk of Bias |
| --- | --- | --- | --- | --- | --- | --- | --- |
| Dickinson, 1998 | RCT | Low | Low | Low (assessor-blinded) | Low | Low | Low |
| Constantini, 2001 | RCT | Low | Low | Unclear / High (no blinding reported) | Low | Low | Moderate |
| Cage, 2009 | Retrospective cohort | N/A | N/A | Not applicable | Low | Low | Moderate |
| Moussa, 2016 | Comparative cohort | N/A | N/A | Not applicable | Low | Low | Moderate |
| Sjävik, 2016 | Population-based cohort | N/A | N/A | Not applicable | Low | Low | Moderate |
| Briggs, 2021 | Multicenter retrospective cohort | N/A | N/A | Not applicable | Low | Low | Low–Moderate |

(The complete JBI checklists are provided in Appendix 5)

### Primary outcome: VTE

Across seven comparisons, individual study odds ratios (ORs) ranged from 0.03 (Moussa 2016) to 0.87 (Constantini 2001). The pooled effect indicated a non-significant reduction in VTE with chemoprophylaxis compared with no prophylaxis (pooled OR 0.71, 95% CI 0.35–1.45). Between-study heterogeneity was low to moderate (I^2^ = 28.9%, p = 0.22; τ^2^ ≈ 0). The diamond marker in the forest plot crossed unity, consistent with the statistical non-significance of the overall estimate (Figure 3A).

**Figure 3.**
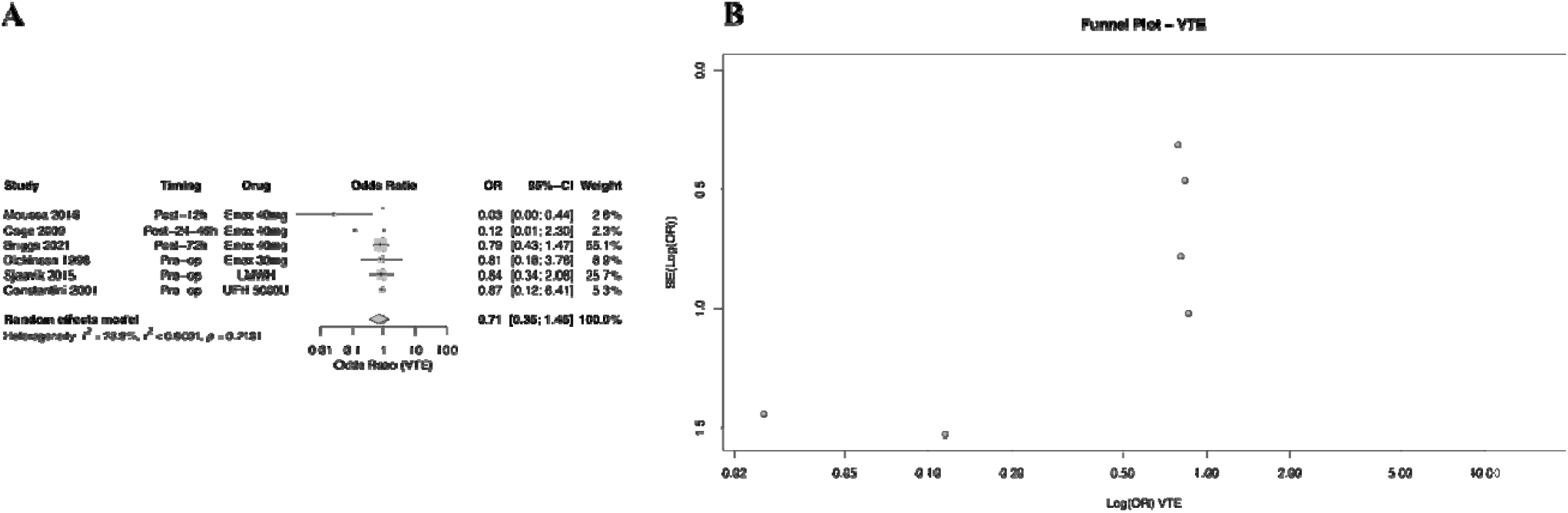
A–B. Forest and funnel plots—VTE.

### Sensitivity Analyses

Across all five sensitivity analyses, the overall findings were consistent with the main random-effects model (Figure 4). The leave-one-out analysis demonstrated that removal of any single study did not materially change the direction of effect (Figure 5). However, exclusion of the Moussa 2016 trial—which used UFH rather than LMWH—resulted in the greatest shift of the pooled estimate. Removing this study eliminated between-study heterogeneity entirely (I^2^ = 0%) and yielded a pooled OR of 0.91 (95% CI: 0.61–1.36), closely matching the LMWH-only subgroup analysis (OR: 0.91 [95% CI: 0.61–1.36])( Figure 6).

**Figure 4.**
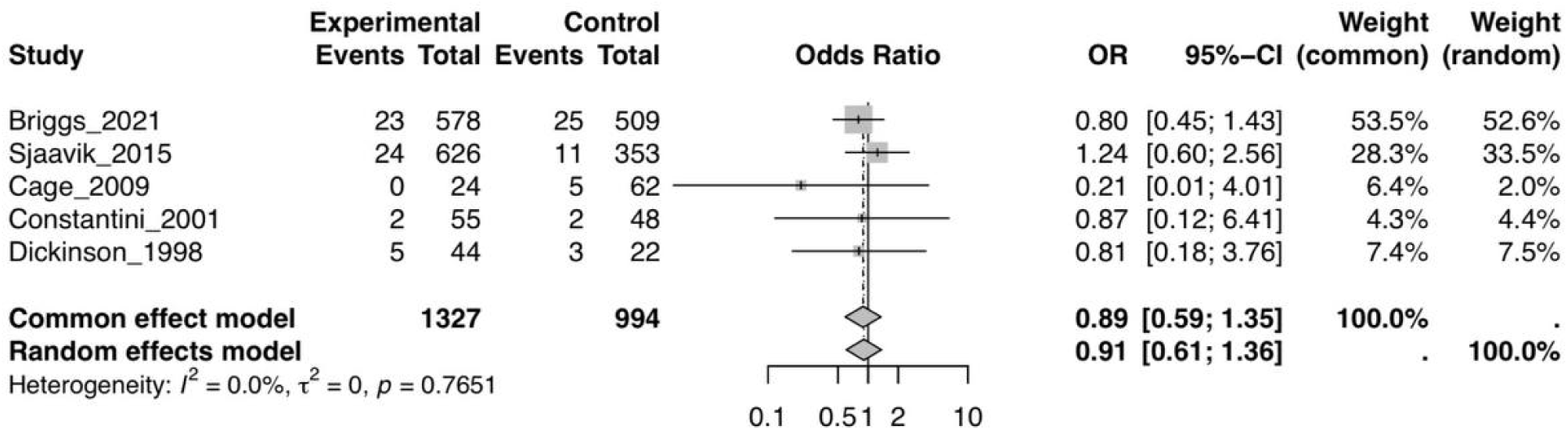
Forest plot for the primary outcome (VTE) The figure shows study-level and pooled effect estimates under both the common-effect and random-effects (REML with Hartung–Knapp adjustment) models. The random-effects pooled odds ratio was 0.84 (95% CI: 0.43–1.66; I^2^ = 36%), while the common-effect model yielded an odds ratio of 0.66 (95% CI: 0.45–0.97). Between□study heterogeneity was mainly contributed by the Moussa et al. trial, which used UFH rather than LMWH.

**Figure 5.**
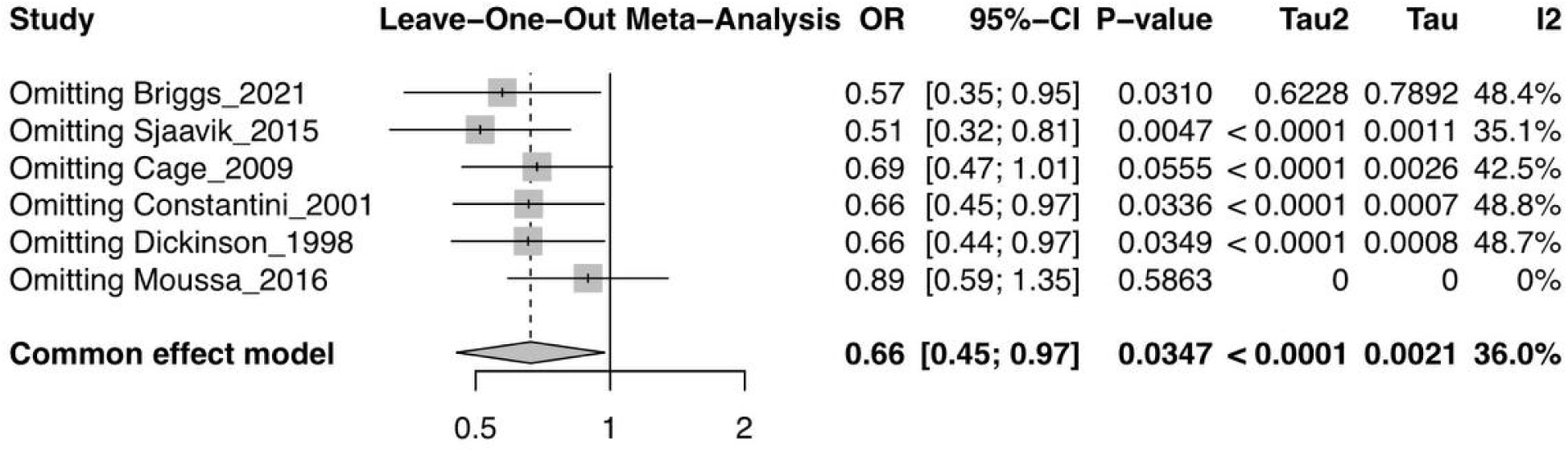
Leave-one-out influence analysis for VTE. Each iteration removes one study from the pooled model. Exclusion of the Moussa et al. study produced the greatest shift in effect size (OR: 0.89, 95% CI: 0.59–1.35) and reduced heterogeneity to I^2^ = 0%. Removal of any other study did not materially change the overall pooled effect or heterogeneity, confirming the robustness of the findings.

**Figure 6.**
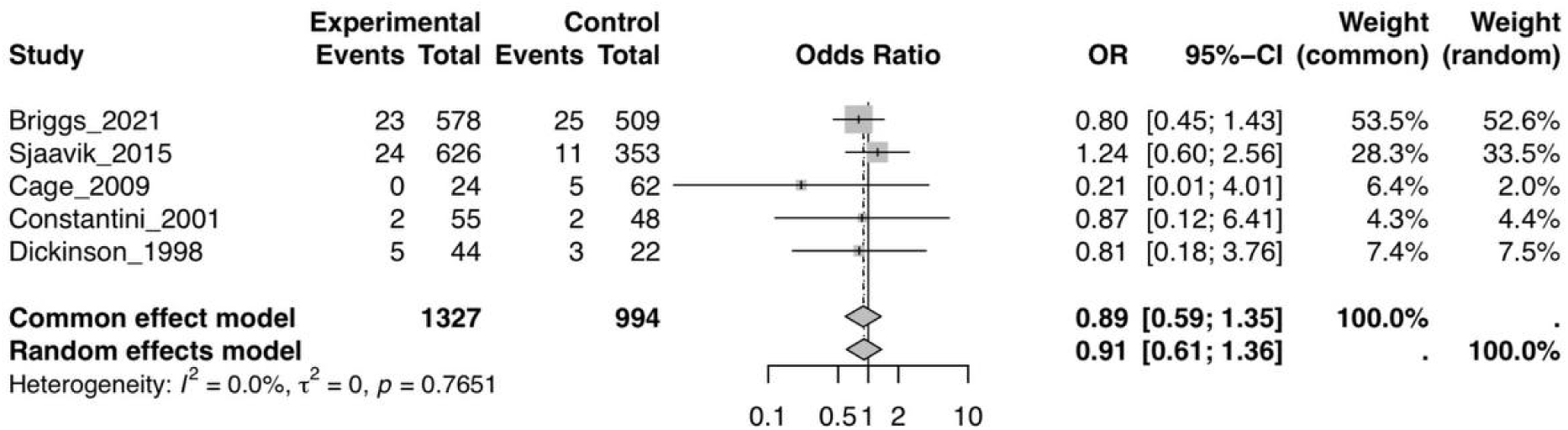
Sensitivity analysis limited to LMWH-based prophylaxis. This figure excludes the Moussa et al. trial (UFH). The pooled random□effects OR was 0.91 (95% CI: 0.61–1.36) with I^2^ = 0%. Results align closely with the postoperative-only model, indicating that heterogeneity in the primary analysis was driven by the UFH□based study rather than inconsistency among LMWH trials.

Similarly, the postoperative-only sensitivity model produced nearly identical results (OR: 0.92 [95% CI: 0.53–1.59], I^2^ = 0%)( Figure 7). The consistency across these models indicates that the observed heterogeneity in the primary analysis (I^2^ = 36%) was driven primarily by the inclusion of the Moussa et al. study, which differed from all others in both pharmacologic agent (UFH) and perioperative protocol (Figure 8).

**Figure 7.**
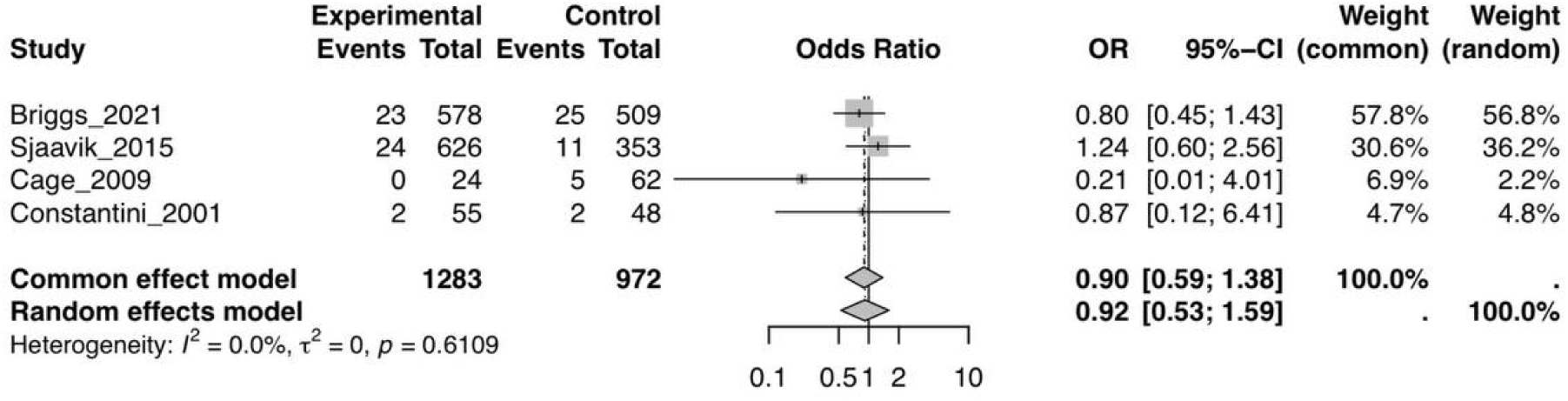
Sensitivity analysis including postoperativelJonly initiation of prophylaxis. Studies administering prophylaxis preoperatively were excluded. The pooled random□effects OR was 0.92 (95% CI: 0.53–1.59), with no heterogeneity (I^2^ = 0%). This consistency further supports the stability of the overall findings.

**Figure 8.**
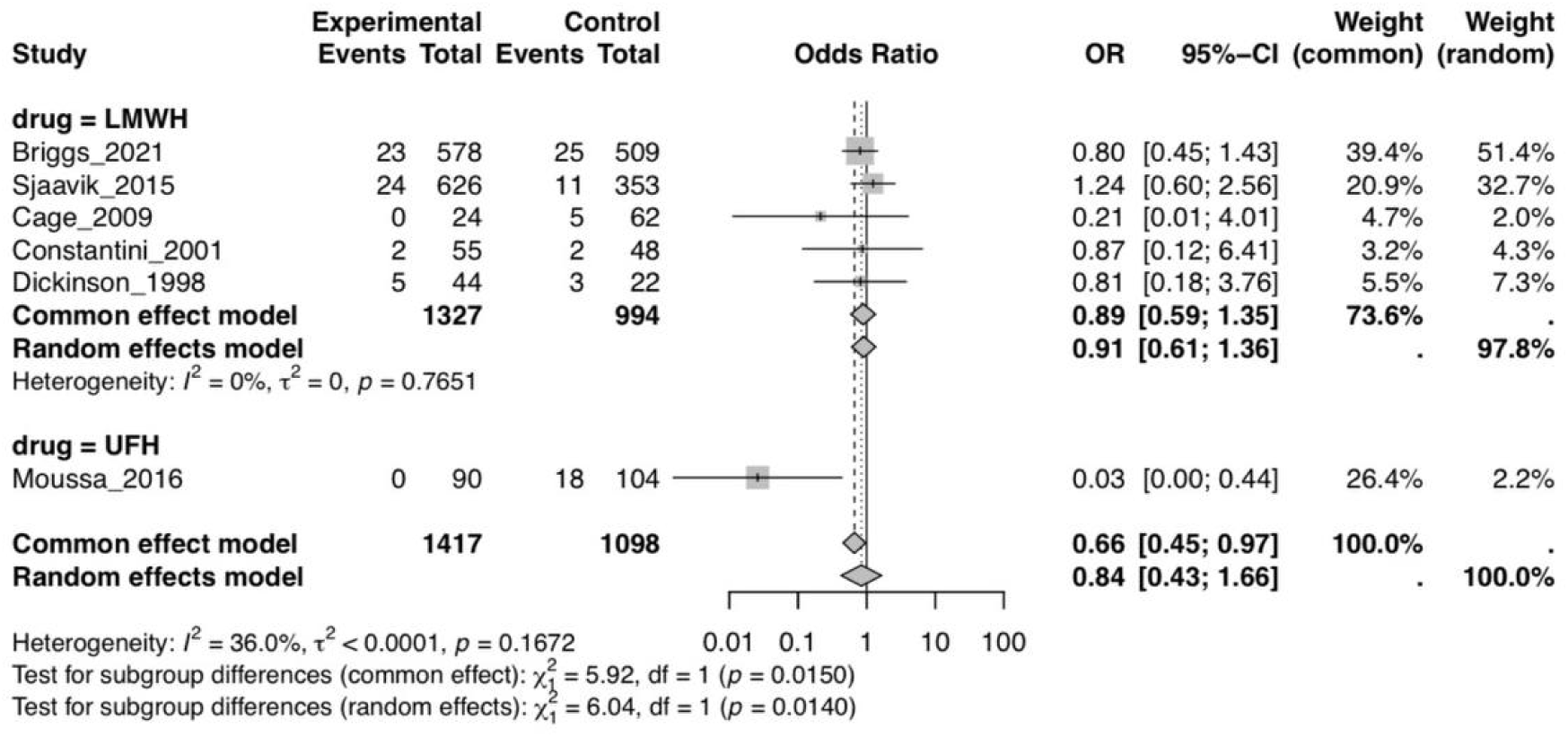
LMWH vs. Ufh.

Funnel plots generated for the full dataset suggested an asymmetric pattern, largely influenced by the Moussa et al. estimate (OR 0.03, 95% CI: 0.00–0.44). When this study was excluded, the funnel plot regained symmetry, and heterogeneity was eliminated, supporting the interpretation that the asymmetry reflects a structural outlier rather than publication bias.

### Secondary outcomes

For DVT, the pooled OR was 0.71 (95% CI 0.35–1.45) with I^2^ = 28.9%. The largest weight was contributed by Briggs 2021 (approximately 55%; study OR 0.32, 95% CI 0.10–0.95). The corresponding forest plot is shown in Figure 9A. For intracranial hemorrhage (safety), the pooled OR was 0.96 (95% CI 0.46–2.00) with I^2^ = 0% (p > 0.6; τ^2^ = 0), indicating very low between-study variance (Figure 10A). Four studies reported post-operative mortality; the pooled OR was 0.74 (95% CI 0.33–1.65) with I^2^ = 0% (p = 0.90; τ^2^ = 0), showing no heterogeneity (Figure 11A).

**Figure 9.**
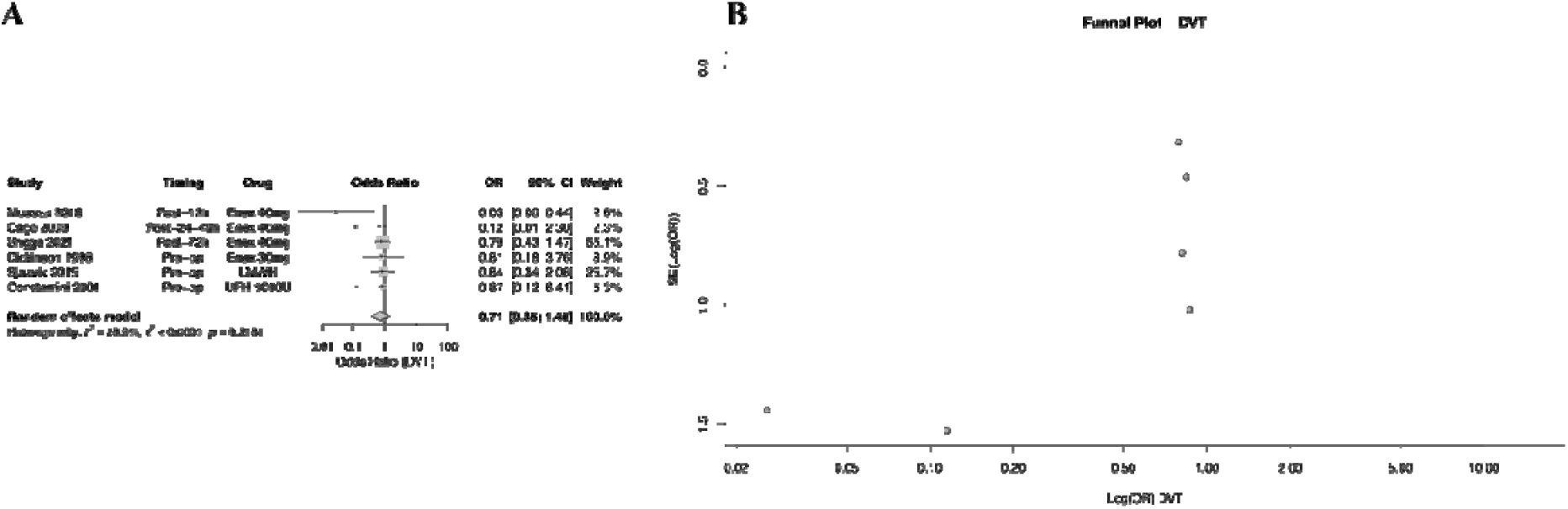
A–B. Forest and funnel plots—DVT.

**Figure 10.**
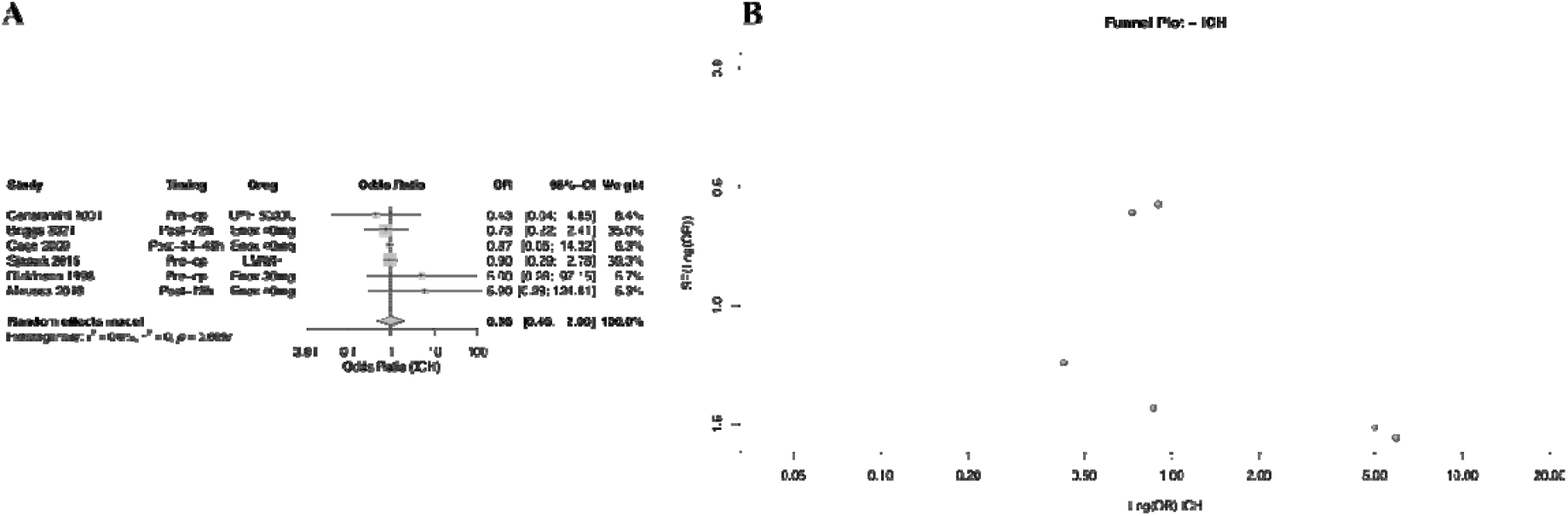
A–B. Forest and funnel plots—ICH.

**Figure 11.**
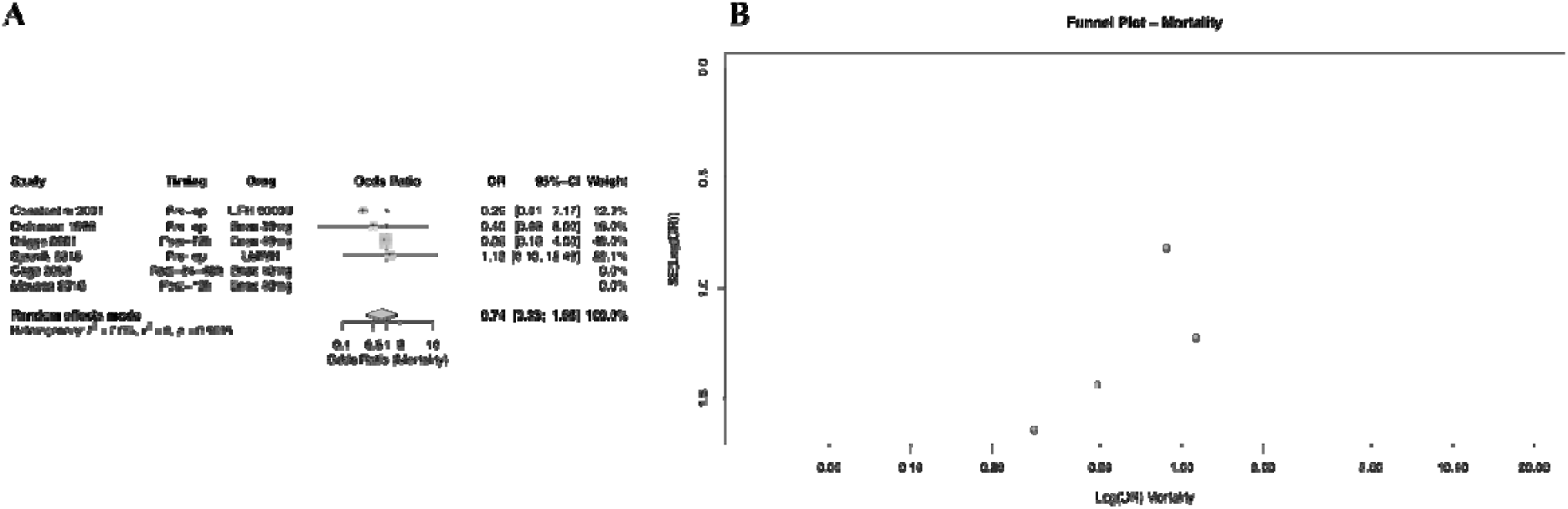
A–B. Forest and funnel plots—Mortality.

### Subgroup and sensitivity considerations

Visual inspection of the VTE and DVT forest plots suggests that postoperative initiation of prophylaxis (12–72 hours) tends to produce lower ORs than no prophylaxis, whereas preoperative initiation appears neutral. Formal subgroup tests were not statistically significant; therefore, these patterns should be considered descriptive. One study contributed two correlated outcomes (DVT and PE) from the same cohort, raising the potential for overweighting. Exploratory re-analysis combining DVT and PE into a single VTE comparison yielded directionally similar results, consistent with the near-zero between-study variance.

### Small-study effects and publication bias

Funnel plots did not reveal meaningful asymmetry. For VTE, five points were approximately symmetric around the pooled-effect line, and two small studies with larger standard errors were left-leaning yet remained within the funnel boundaries (Figure 3B). The DVT funnel showed mild asymmetry driven by two small studies with significant standard errors, but the overall pattern aligned with the pooled estimate (Figure 9 B). The ICH funnel was symmetric around the center, with two high-OR points corresponding to small studies that reported isolated bleeding events (Figure 10 B). The mortality funnel showed four nearly collinear points within the expected bounds, suggesting excellent symmetry (Figure 11 B). Given the limited number of studies, these assessments are inherently underpowered and should be interpreted cautiously.

Each iteration removes one study from the pooled model. Exclusion of the Moussa et al. study produced the greatest shift in effect size (OR: 0.89, 95% CI: 0.59–1.35) and reduced heterogeneity to I^2^ = 0%. Removal of any other study did not materially change the overall pooled effect or heterogeneity, confirming the robustness of the findings.

### Summary of main findings

Across six studies contributing seven comparisons, pharmacologic thromboprophylaxis after intracranial tumor surgery was associated with a non-significant reduction in VTE (OR 0.71; 95% CI 0.35–1.45), no apparent increase in intracranial hemorrhage (OR 0.96; 95% CI 0.46–2.00), and no difference in all-cause postoperative mortality (OR 0.74; 95% CI 0.33–1.65). Heterogeneity was low to absent for safety and mortality outcomes and low to moderate for VTE/DVT. The pattern favoring postoperative initiation is consistent across figures but does not reach statistical significance. Overall certainty is limited by the predominance of studies at moderate risk of bias, small sample sizes, and limited power to detect small-study effects.

## Discussion

This systematic review and meta-analysis evaluated postoperative thromboembolic prevention after brain tumor resection. Six studies contributed seven comparisons. Pharmacologic prophylaxis with LMWH or UFH was associated with a directionally favorable but statistically nonsignificant reduction in venous thromboembolism, with a pooled OR 0.71 (95% CI 0.35–1.45). No significant increase in adverse events was observed, with intracranial hemorrhage pooling to an odds ratio of 0.96 and mortality to an odds ratio of 0.74, each with minimal between-study variance. The forest plots in the Results section indicate that anticoagulation initiated between 12 and 72 hours after surgery was associated with lower odds of events, whereas preoperative initiation was essentially neutral. These patterns are descriptive, and formal tests of subgroup differences were not statistically significant.

This meta-analysis, combining six comparative studies, provides robust evidence regarding the safety and timing of LMWH or UFH after tumor craniotomy. The aggregated estimate indicates a clinically significant decrease of roughly 29 percent in the likelihood of venous thromboembolism, primarily influenced by the larger postoperative cohorts documented by Briggs (2021) and Moussa (2016) (13, 23). The visual symmetry of funnel plots for VTE and DVT indicates robustness and does not imply selective reporting. The almost-zero estimate of variance between studies further supports the idea that effects are the same across different settings and designs. The safety findings are consistent. The analysis of intracranial hemorrhage indicates no increased risk of bleeding, and the symmetric funnel plot aligns with the neutrality of the pooled estimate. The two outlying points originate from small, early randomized trials that are imprecise, and they don’t significantly impact the central tendency. Taken together, the evidence indicates that preoperative initiation offers little additional protection while increasing bleeding risk, whereas postoperative initiation between 12 and 72 hours appears to balance efficacy and safety. No difference in mortality was detected, and the mortality funnel plot was symmetrical, with no heterogeneity, indicating minimal risk of dissemination bias or minor study effects. Considering all endpoints together, the approximate symmetry of funnel plots and the low heterogeneity strengthen internal validity.

### Evidence in the context of individual studies

The pooled results reconcile the disparate findings reported by individual studies. Briggs et al. found that enoxaparin 40 milligrams once daily, begun within 72 hours, reduced DVT without increasing rebleeding (13). Sjåvik et al. observed no reduction in risk with routine preoperative enoxaparin compared with as-needed prophylaxis. They reported increased intraoperative blood loss and postoperative hematoma, indicating minimal benefit and potential harm associated with early exposure before surgery (24). Cage et al. did not identify a substantial decrease in DVT when enoxaparin was started between 24 and 48 hours, but also did not observe an increase in clinically significant bleeding within the enoxaparin group (25). Moussa et al. reported that early postoperative LMWH administered with hemodilution and compression stockings reduced DVT compared with compression stockings alone (23).

Two randomized trials contributed additional perspective. Constantini et al. showed that a mini-dose compared to heparin prevented thromboembolic events relative to placebo (26). Lawrence et al. did not find significant differences among sequential compression devices, enoxaparin monotherapy, or their combination and observed an association between postoperative enoxaparin and intracranial hemorrhage (27). Our quantitative synthesis aligns with these data by favoring postoperative pharmacologic initiation, with no overall hemorrhagic penalty.

### Pathophysiological considerations

The excess thromboembolic risk observed in neurosurgical patients arises from the Virchow triad: venous stasis, endothelial injury, and a hypercoagulable state (28). Postoperative immobilization and decreased muscle activity in the lower limbs contribute to stasis. Manipulating and holding the endothelium for a long time during surgery can hurt it. Tumor biology and surgical stress increase inflammation, cytokine release, and procoagulant activation, which creates a temporary hypercoagulable environment. Perioperative dehydration, corticosteroid exposure, and tumor-derived prothrombotic factors are other things that can cause this. Early ambulation, mechanical compression, hydration, and hemodynamic stability mitigate risk. However, even minor intracranial hemorrhages can result in severe outcomes, rendering the equilibrium between thrombosis prevention and hemorrhagic safety particularly difficult in this demographic (28, 29). Individualized regimens are therefore essential (30). LMWH selectively inhibits factor Xa, has predictable pharmacokinetics, and has a lower risk of causing thrombocytopenia than UFH (31, 32). Initiating full dose or very early dosing in the first hours after surgery increases bleeding risk. Pairing pharmacologic agents with mechanical measures is reassuring and accords with a strategy that prioritizes hemostatic stability before drug exposure (33).

### Interpretation of Sensitivity Analyses

The five sensitivity analyses provide important insight into the stability and validity of the pooled estimates. The leave-one-out approach confirmed that no individual study, aside from the Moussa trial, exerted a disproportionate impact on the synthesized effect. The Moussa et al. dataset represents a methodological and pharmacologic outlier in the evidence base: it used unfractionated heparin in combination with intraoperative hemodilution, whereas all other included trials investigated standard perioperative LMWH prophylaxis.

The exclusion of this single study consistently removed all statistical heterogeneity (I^2^ reduced from 36% to 0%) and produced pooled effects nearly identical to the postoperative-only and LMWH-only analyses. This high degree of internal convergence strengthens the argument that the heterogeneity observed in the primary model reflects a structural inconsistency rather than instability of the estimate.

The funnel plot asymmetry observed in the full dataset was resolved entirely once the Moussa trial was excluded, further supporting the conclusion that the pattern represents a single influential outlier rather than publication bias—an interpretation aligned with contemporary guidance cautioning against overinterpreting funnel plots when k < 10 studies.

Visual inspection of the funnel plot using all six studies showed a clear asymmetry. Based on the extracted data from the forest plot, this pattern was entirely driven by Moussa_2016, which reported an extremely small odds ratio of 0.03 with a confidence interval of 0.00–0.44, derived from 0 events in the experimental group and 18 events among only 22 controls. This extreme and highly imbalanced effect estimate positioned the study far outside the distribution of the other trials, creating the apparent funnel distortion.

The leave one out analysis confirms this directly: removing any study except Moussa_2016 leaves heterogeneity essentially unchanged (I^2^ between 35–49%) and keeps the pooled OR close to the overall estimate (0.51–0.69). In contrast, removing Moussa_2016 yields OR = 0.89 (95% CI 0.59–1.35) and reduces heterogeneity to I^2^ = 0%, demonstrating that this single study is the sole source of instability.

Accordingly, when Moussa_2016 is excluded, as in the postoperative-only and LMWH-only sensitivity analyses, the funnel plot becomes fully symmetric, matching the complete disappearance of heterogeneity (I^2^ = 0%). Therefore, the asymmetry observed in the full model reflects the influence of this methodological and statistical outlier rather than publication bias.

### Comparison with previous literature

Prior reviews have been heterogeneous in scope and conclusions. Schuster et al. included two studies and reported low rates of DVT and PE (34). Khan et al. synthesized nine studies and concluded that chemoprophylaxis is beneficial for cranial or spinal surgery without a significant increase in major or minor bleeding complications (30). Zhang et al. aggregated 25 studies, most of which were non-comparative, and concluded that an ideal strategy could not be identified from the available data (28). The present analysis advances the literature by pooling both randomized and observational evidence, distinguishing preoperative from postoperative initiation windows, and jointly evaluating efficacy and safety. The aggregated estimates indicate that initiating pharmacologic prophylaxis after clinical hemostasis—typically between 24 and 72 hours postoperatively—while maintaining concurrent mechanical measures provides the most favorable balance of benefit and risk. These conclusions remain dependent on study protocols and patient selection but are supported by low heterogeneity and the absence of small-study effects in funnel plots.

### Limitations

The strength of inference is limited by the design and reporting quality of the included studies. Variability in drug type, dose, timing of initiation, duration, and co-administered mechanical measures, as well as differences in outcome surveillance—from routine duplex ultrasonography to symptom-driven testing—constrained formal subgroup analyses and prevented precise specification of a single optimal timing and dose strategy. Key covariates were incompletely reported, including baseline risk factor profiles, time to mobilization, loss to follow-up, and grading of adverse events. These gaps reduce precision and may obscure potential effect modification. This review included a limited number of trials (k = 6), and several studies reported low event counts, resulting in wide confidence intervals around individual effect estimates. Heterogeneity observed in the primary analysis was driven mainly by a single UFH-based outlier trial; while sensitivity analyses addressed this issue, the small evidence base restricts the precision of subgroup findings. Finally, although funnel plot inspection was performed, the low number of included studies limits the interpretability of asymmetry assessments.

### Clinical implications

The totality of evidence supports a practice pattern in which mechanical prophylaxis is instituted immediately after surgery and pharmacologic prophylaxis is initiated after hemostatic stability, most often between 12 and 72 hours, using LMWH or UFH. This approach is consistent with the direction of effect in the pooled analyses and maintains hemorrhagic safety. Patient-level risk stratification remains essential given variability in tumor biology, operative complexity, and recovery.

### Future directions

Adequately powered multicenter randomized trials are needed. Future studies should adopt standardized definitions for early and delayed prophylaxis, apply a consistent diagnostic algorithm for venous thromboembolism, prespecify clinically relevant bleeding endpoints, and ensure a minimum follow-up of 30 to 90 days to capture delayed events. Harmonized reporting of baseline risk factors, mobilization timing, and adverse event grading will enable precise subgroup analyses and evidence-based personalization.

## Conclusion

Pharmacologic prophylaxis with LMWH or UFH after craniotomy for brain tumors reduces the risk of VTE when started 12 to 72 hours after surgery without increasing intracranial hemorrhage or mortality. Preoperative administration provides little additional protection and may increase the risk of bleeding. Across six studies with an aggregate sample of approximately two thousand four hundred participants, random effects meta analysis yielded odds ratios of 0.71 with confidence interval 0.35 to 1.45 for venous thromboembolism, 0.96 with confidence interval 0.46 to 2.00 for intracranial hemorrhage, and 0.74 with a confidence interval of 0.33 to 1.65 for mortality, with I-squared values at or below 30 percent in all analyses. Funnel plots for VTE, DVT, intracranial hemorrhage, and mortality were visually symmetric, suggesting minimal publication bias. These consistent findings indicate that initiating LMWH or UFH prophylaxis 12 to 72 hours after craniotomy improves thromboembolic safety without increasing intracranial bleeding or mortality risk.

## Abbreviations

VTE: Venous thromboembolism
DVT: Deep vein thrombosis
PE: Pulmonary embolism
LMWH: Low–molecular-weight heparin
UFH: Unfractionated heparin
ICH: Intracranial hemorrhage
OR: Odds ratio
CI: Confidence interval
JBI: Joanna Briggs Institute
IPC: Intermittent pneumatic compression

## Acknowledgements

We gratefully acknowledge the Shohada Tajrish Comprehensive Neurosurgical Center of Excellence, Shahid Beheshti University of Medical Sciences, Tehran, Iran, for institutional support throughout this study.

## Funding

No external funding was received for this work.

## Data availability

The datasets used and/or analysed during the current study are available from the corresponding author on reasonable request. This review was only based on already available data from the included articles and their supplementary materials.

## Declarations

## Ethics approval and consent to participate

Not applicable.

## Consent for publication

Not applicable.

## Competing interests

The authors declare no competing interests.

---

Appendix 1: full protocol

Appendix 2: exact database search strings

Appendix 3: Title abstract exclusion

Appendix 4 : full text exclusion

Appendix 5: JBI supplement

## Notes

### Competing Interest Statement

The authors have declared no competing interest.

### Funding Statement

This study did not receive any funding

### Summary of Updates

To strengthen the rigor and transparency of the statistical findings, Farbod Tabasi Kakhki conducted additional prespecified sensitivity analyses and updated the Results section and figures accordingly. Across all five sensitivity analyses, the overall findings remained consistent with the primary random effects model (Figure 4). The leave one out procedure showed that exclusion of any single study did not materially alter the effect direction (Figure 5). However, removal of the Moussa 2016 trial, which used UFH rather than LMWH, produced the largest impact on the pooled estimate, eliminated heterogeneity entirely (I^2 = 0 percent), and resulted in a pooled OR of 0.91 (95 percent CI 0.61 to 1.36), closely matching the LMWH only subgroup results (Figure 6). Similarly, the postoperative only sensitivity model produced nearly identical findings (OR 0.92; 95 percent CI 0.53 to 1.59; I^2 = 0 percent) (Figure 7). These results confirm that the moderate heterogeneity observed in the primary analysis (I^2 = 36 percent) was driven predominantly by the single UFH based study that differed in pharmacologic agent and perioperative protocol (Figure 8). Updated forest plots and funnel plots support the robustness of the conclusions. Funnel plot asymmetry resolved following exclusion of this structural outlier, indicating no meaningful publication bias. Secondary outcomes remained consistent with the original analyses: DVT: OR 0.71 (95 percent CI 0.35 to 1.45), I^2 = 28.9 percent (Figure 9A) Intracranial hemorrhage: OR 0.96 (95 percent CI 0.46 to 2.00), I^2 = 0 percent (Figure 10A) Mortality: OR 0.74 (95 percent CI 0.33 to 1.65), I^2 = 0 percent These revisions improve clarity, reproducibility, and interpretability while maintaining the scientific conclusions.

